# COVID-19, Type-2 Diabetes, and Associated Health Outcomes in China: Results from a Nationwide Survey of 10,545 Adults

**DOI:** 10.1101/2020.10.07.20207282

**Authors:** Zumin Shi, Alice Yan, Paul Zimmet, Xiaoming Sun, Nayla Cristina do Vale Moreira, Lawrence J. Cheskin, Liming Wang, Weidong Qu, Hong Yan, Akhtar Hussain, Youfa Wang

**Affiliations:** Human Nutrition Department, College of Health Sciences, QU Health, Qatar University, Doha, Qatar; Center for Advancing Population Science, Division of General Internal Medicine, Department of Medicine, Medical College of Wisconsin, Milwaukee, WI, USA; Department of Diabetes, Central Clinical School, Monash University, Melbourne, Australia; Global Health Institute, School of Public Health, Xi’an Jiaotong University Health Science Center, Xi’an, China; Faculty of Medicine, Federal University of Ceará (FAMED-UFC), Brazil; Institute of Health and Society, Department of Community Medicine and Global Health, University of Oslo, Oslo, Norway; Department of Nutrition and Food Studies, George Mason University, Fairfax, VA 220030, USA; National Center for Chronic and Non-communicable Disease Control and Prevention, Chinese Center for Disease Control and Prevention, Beijing, China; Centers for Water and Health, Key Laboratory of the Public Health Safety, Ministry of Education, Department of Environmental Health, School of Public Health, Fudan University, Shanghai, 200032, China; Faculty of Health Sciences, Nord University, Bodø 8049, Norway; International Diabetes Federation. 166 Chaussee de La Hulpe B-1170, Brussels, Belgium

**Keywords:** COVID-19, pandemic, diabetes, non-communicable diseases, health outcomes, China

## Abstract

**Objective:** This study examined the associations between type-2 diabetes (T2DM) and self-reported/familial COVID-19 infection and investigated health-related outcomes among those with diabetes during China’s nationwide quarantine.

**Methods:** The 2020 China COVID-19 Survey was administered anonymously via social media (WeChat) across China. It was completed by 10,545 adults in all of mainland China’s 31 provinces. The survey consisted of 74 items covering sociodemographic characteristics, preventive measures for COVID-19, lifestyle behaviors, and health-related outcomes during the period of quarantine. Regression models examined associations among study variables, adjusting for covariates.

**Results:** Diabetes was associated with a six-fold increased risk of reporting COVID-19 infection among respondents or their family members. Among patients with diabetes, individuals who rarely wore masks had double the risk of suspected COVID-19 infection compared with those who always wore masks, with an inverse J-shaped relationship between face mask wearing and suspected COVID-19 infection. People with T2DM tended to have both poor knowledge of COVID-19 and poor compliance with preventive measures, despite perceiving a high risk of personal infection (40.0% among respondents reporting T2DM and 8.0% without T2DM). Only 54-55% of these respondents claimed to consistently practice preventive measures, including wearing face masks. Almost 60% of those with T2DM experienced food or medication shortages during the quarantine period, which was much higher than those without T2DM. Importantly, respondents who experienced medication shortages reported a 63% higher COVID-19 infection rate.

**Conclusions:** T2DM was associated with an increased risk of self-reported personal and family member COVID-19 infection, which is mitigated by consistent use of face masks.

**Funding:** The project is supported in part by research grants from the China Medical Board (Grant number: 16-262), the National Key Research and Development Program of China (Grant Number: 2017YFC0907200 & 2017YFC0907201), the University Alliance of the Silk Road (Grant number: 2020LMZX002), and Xi’an Jiaotong University Global Health Institute.

**Research in Context:** *Evidence before this study:* During the COVID-19 pandemic, it has become increasingly clear that the risk factors for initial infection and subsequent poor health outcomes include, but are not limited to, social vulnerability, economic status, older age, and obesity. While community-wide masking has been recommended by the World Health Organization to control COVID-19, its overall effectiveness has not been clearly evaluated.

*Added value of this study:* Through an anonymous survey disseminated and promoted through WeChat, the largest social media platform in China, we sought to understand the impact of COVID-19 on the health, wellbeing, and health-related behaviors of adults in China. Specifically, this study examined how individuals with chronic diseases managed the threat, including their COVID-19 related knowledge, attitudes, and adherence to preventive measures such as wearing face masks, and their disease-related self-care.

*Implications of the available evidence:* This study demonstrates that type-2 diabetes mellitus is associated with an increased risk of COVID-19 infection, which is mitigated by consistent use of face masks.

## INTRODUCTION

The COVID-19 pandemic has become a major concern worldwide. Diabetes mellitus (DM) has been identified as one of the most commonly reported non-communicable diseases (NCDs) associated with COVID-19.^1^ People with DM, especially type 2 diabetes (T2DM), when infected with SARS-CoV-2 are susceptible to worse clinical outcomes (higher hospitalization and mortality rates).^1^ In addition to impaired immunity, increased coagulation, and chronic inflammation,^2^ several other factors have been identified that impact the severity of COVID-19 among people with diabetes – including poor glycemic control, the pharmacodynamics of some medications commonly used in diabetes care, and limited access to medications.^3,4^

The COVID-19 outbreak was first reported in China, followed by a nationwide lockdown. Several COVID-19 related prevention and control measures, including social distancing and quarantine practices can result in decreased physical activity, poor dietary diversity, and psychological distress, well as delayed care-seeking related to fear of exposure to COVID-19.^5,6^. Given that China has the largest number of people with diabetes in the world (an estimated 116 million adults),^7^ it would be critical to understand how Chinese patients with diabetes mitigate the risk of COVID-19, and manage their diabetes.

To control the spread of the virus, studies have highlighted the importance of individual adherence to preventive measures, such as use of face masks, frequent handwashing, proper respiratory hygiene, and social distancing.^8^ An online cross-sectional survey from China with 6,910 subjects reported that most participants engaged in appropriate preventive practices, especially wearing masks and avoiding crowded places. Furthermore, higher levels of knowledge of COVID-19 was associated with a greater likelihood of compliance with preventive measures.^9^ However, this study had limited sample representativeness, with an over-representation of women and well-educated people in only one province in China.

So far, little is known about the psychosocial and health-related consequences that the pandemic has had on adults with diabetes.^10^ Considering the lack of nationally-available data on public health measures and responses, the level of knowledge of COVID-19 and adherence to preventive measures among the Chinese population, especially those with diabetes, remain open questions.

Using nationwide survey data from China, this study investigated: 1) the association between diabetes and self-reported COVID-19 infection in families; 2) knowledge of and practices of people towards mitigating and controlling the spread of COVID-19, and 3) the psychosocial and health-related impact of the COVID-19 pandemic on those with diabetes during the national quarantine. The findings may be useful in guiding prevention and control of COVID-19 as well as the management of T2DM during the pandemic.

## METHODS AND MATERIALS

### Study design and participants

The 2020 China COVID-19 Survey is a cross-sectional, anonymous online survey that was administered via WeChat between April 25 and May 11, 2020. We selected this platform not only because the nation was under quarantine and we could only reach respondents online, but also because WeChat is China’s leading social network, with more than one billion users, and the majority of Chinese adults use WeChat daily. Based on national usage statistics, WeChat’s penetration rate is 78% in China among 16-64-year-old in 2020.^11^ To recruit a diverse sample of respondents, we used both snowball and convenience sampling approaches.

The survey questionnaire contained 74 items, provides about 150 study variables, and covered 8 topics, namely: 1) knowledge, attitudes, beliefs, and practices associated with COVID-19; 2) personal experiences and impacts of COVID-19; 3) attitude toward government responses to COVID-19; 4) healthcare-seeking behaviors; 5) demographic characteristics; 6) lifestyle behaviors; 7) psychological well-being; and 8) health outcomes, including obesity and other chronic diseases during the nationwide quarantine.

The study was approved by the Xian Jiaotong University’s Institutional Review Board, and participants provided consent. Data includes a nationwide (31 province-level administrative units) sample of 10,545 adults aged ≥18 years.

### Measures

#### Outcome measure: Self-reported COVID-19 infection and health-related outcomes

##### COVID-19 infection in the family

Participants were asked three yes-no questions: 1. “Do you think that you or someone in your family have been infected with COVID-19?” We defined those who answered in the affirmative as having self-reported suspected COVID-19 infection. 2. “Have you visited the hospital due to suspected COVID-19 infection?” 3. Discriminatory behavior related to COVID-19 was assessed through the question, “Have you or anyone in your family experienced COVID-19-related bias or discrimination due to infection with COVID-19?” Notably, we defined “possible COVID-19 infection” only in those reporting a positive answer to all three questions above.

##### Self-reported health worsening

Participants were asked, “How has COVID-19 affected your health? 1) Significantly worse; 2) Slightly worse; 3) Slightly better; 4) Significantly better; or 5) No change.”

*Weight change* during the nationwide lockdown was gauged by the question “How is your current weight compared to your weight before COVID-19? 1) Unchanged (increased or decreased by less than 1 kg); 2) Increased by 1.0 to 2.5 kg; 3) Increased by 2.5 kg or more; 4) Decreased by 1.0-2.5 kg; or 5) Decreased by 2.5 kg or more.”

##### Worry index

Participants were asked five questions about their “stressful experiences” during the pandemic in the month prior to the survey: 1) loss of interest in activities they liked in the past, 2) difficulty falling asleep, staying asleep, or waking up frequently or early, 3) irritability or anger, 4) difficulty concentrating, and 5) repeated disturbing dreams associated with COVID-19. Response options for each question included a five-point Likert scale from “not at all” to “extremely.” These questions were adopted from diagnostic screening items of the widely used and validated PCL-C (civilian) scale,^12^ which asks about symptoms in relation to “stressful experiences” and can be used in any population.

A worry index was constructed based on responses to the five questions. Each question had a score between 1 to 5 (not bothered to extremely bothered). We calculated the total score (range of 5-25) and then converted it to a z-score.

#### Diabetes and other NCDs

To assess NCDs, participants were asked to respond to the following questions: “Do you currently have a chronic disease?” If so, they were asked to identify specific diseases or health conditions. In the analysis, we categorized participants as “Without diabetes, with diabetes, and with other NCDs (including hypertension, heart disease, stroke, cancer, asthma, chronic lung disease, et al).”

#### Knowledge, attitudes, and practices towards COVID-19

These were assessed using items adapted from a recent COVID-19 awareness, attitude, and action questionnaire.^13^ Two of the questions asked participants to rate their perceived likelihood that they, or a family member, would become infected with COVID-19, and levels of worry – both on a four-point scale, from 1 “not at all likely/not at all worried” to 4 “very likely/very worried.” Participants were also asked how they complied with the four main preventive measures (wearing a face mask, frequent hand washing, avoiding going out, and avoiding social gatherings) on a five-point Likert scale (all the time, sometimes, occasionally, rarely, and never).

#### Health-related experiences and behavior changes during COVID-19

We adopted a questionnaire developed by Conway and colleagues (2020) to measure the impact and personal experiences related to COVID-19.^14^ Two items – medication shortage and food shortage due to COVID-19 – were used to measure coronavirus-related experiences. Body mass index (BMI) was calculated using self-reported weight and height. We used BMI ≥24 kg/m^2^ to define overweight/obese. Physical activity during COVID-19 was measured with items adapted from the International Physical Activity Questionnaire (IPAQ).^15^ We then calculated participants’ weekly physical activity minutes and whether they met the 150 minute weekly physical activity recommendation. Participants’ self-rated health^16^ was measured by the most commonly used item from the Patient-Reported Outcomes Measurement Information System (PROMIS), with responses ranging from “poor” to “excellent.”

#### Demographic variables

Demographic characteristics include gender, education, income, and residence (city, town, or rural) and were included as covariates in analysis models.

### Statistical analysis

The sample characteristics were presented as means (SD) or proportions. To compare differences between groups, we used a X^2^ test for categorical variables, and ANOVA for continuous variables.

We used bar charts to graphically present the distributions of practicing preventive measures for COVID-19. Multivariable Poisson regression with robust variance was used to calculate prevalence ratios (PR),^17^ and assess the association between diabetes and significant health worsening during the pandemic while controlling for sociodemographic variables. It is usually preferable to use PRs instead of odds ratios (ORs) in cross-sectional studies when the prevalence of outcome measures is above 10%.

To examine the association between diabetes status and worry index, we employed multivariable linear regression. Multivariable logistic regression was used to assess the association between diabetes and self-reported COVID-19 infection. We conducted and visually presented our subgroup analyses of the association between diabetes and self-reported COVID-19 infection. Among those reporting diabetes, the association between sociodemographic factors and facemask wearing with self-reported COVID-19 infection was examined using Poisson regression with robust variance, as the prevalence of self-reported COVID-19 was above 10%.

All analyses were performed using STATA 16.1 (College Station, TX, USA). Statistical significance was considered when p<0.05 (two-sided).

## RESULTS

### Sample characteristics

**Table 1** shows the sample characteristics by diabetes status. Among the 10,545 participants (4,605 men and 5,940 women) 585 (6.5%) reported having T2DM and 1,529 (14.5%) had other NCDs. Compared to participants without diabetes or with other NCDs, more participants with diabetes were men (60.3%) and were older. They were more likely to live in cities (68.7%), had a lower level of education, a higher level of income (47.4%), and were more likely to be overweight (23.8%) or obese (10%). Compared to those without diabetes, those with diabetes were more likely to be current smokers (43.6% vs. 11.8%, p<0.001) and current alcohol drinkers (40.2% vs. 19.7%, p<0.001). About 60% of those with diabetes reported experiencing food (59.8%) or medication (59.7%) shortage, which was much higher than those without diabetes (22.7% and 25.8%).

**Table 1.** Sample characteristics among participants attending “The 2020 China COVID 19 Survey”, by diabetic and non-communicable chronic disease (NCD) status (N=10,545) (77.3%)

|  | **Total**<br>**N=10,545** | **Without**<br>**diabetes**<br>**N=8,431** | **With**<br>**diabetes**<br>**N=585** | **With other**<br>**NCD**<br>**N=1,529** | **p-**<br>**value** |
| --- | --- | --- | --- | --- | --- |
| Age group (years) |  |  |  |  | <0.001 |
| 18-29 | 4,989<br>(47.3%) | 4,119 (48.9%) | 288 (49.2%) | 582 (38.1%) |  |
| 30-39 | 3,493<br>(33.1%) | 2,844 (33.7%) | 167 (28.5%) | 482 (31.5%) |  |
| 40-49 | 1,352<br>(12.8%) | 1,016 (12.1%) | 66 (11.3%) | 270 (17.7%) |  |
| 50-59 | 592 (5.6%) | 388 (4.6%) | 49 (8.4%) | 155 (10.1%) |  |
| 60-80 | 119 (1.1%) | 64 (0.8%) | 15 (2.6%) | 40 (2.6%) |  |
| Sex |  |  |  |  | <0.001 |
| Men | 4,605<br>(43.7%) | 3,486 (41.3%) | 353 (60.3%) | 766 (50.1%) |  |
| Women | 5,940<br>(56.3%) | 4,945 (58.7%) | 232 (39.7%) | 763 (49.9%) |  |
| Education |  |  |  |  | <0.001 |
| Low (primary school or lower) | 191 (1.8%) | 95 (1.1%) | 37 (6.3%) | 59 (3.9%) |  |
| Medium (junior high/high school) | 3,985<br>(37.8%) | 2,924 (34.7%) | 280 (47.9%) | 781 (51.1%) |  |
| High (university or above) | 6,369<br>(60.4%) | 5,412 (64.2%) | 268 (45.8%) | 689 (45.1%) |  |
| Residence |  |  |  |  | <0.001 |
| City | 6,493<br>(61.6%) | 5,148 (61.1%) | 402 (68.7%) | 943 (61.7%) |  |
| Town | 2,470<br>(23.4%) | 1,980 (23.5%) | 96 (16.4%) | 394 (25.8%) |  |
| Rural | 1,582<br>(15.0%) | 1,303 (15.5%) | 87 (14.9%) | 192 (12.6%) |  |
| Income levels (tertiles) |  |  |  |  | <0.001 |
| Low | 3,592<br>(42.5%) | 2,920 (43.9%) | 161 (32.7%) | 511 (39.3%) |  |
| Medium | 2,152<br>(25.5%) | 1,748 (26.3%) | 98 (19.9%) | 306 (23.6%) |  |
| High | 2,704<br>(32.0%) | 1,989 (29.9%) | 233 (47.4%) | 482 (37.1%) |  |
| Occupation |  |  |  |  | <0.001 |
| Student | 2,075<br>(19.7%) | 1,766 (20.9%) | 110 (18.8%) | 199 (13.0%) |  |
| Employed | 7,015<br>(66.5%) | 5,630 (66.8%) | 367 (62.7%) | 1,018 (66.6%) |  |
| Unemployed | 1,135<br>(10.8%) | 854 (10.1%) | 76 (13.0%) | 205 (13.4%) |  |
| Retired | 320 (3.0%) | 181 (2.1%) | 32 (5.5%) | 107 (7.0%) |  |
| Smoking |  |  |  |  | <0.001 |
| Current smoker | 1,633<br>(15.5%) | 993 (11.8%) | 255 (43.6%) | 385 (25.2%) |  |
| None smoker | 8,148<br>(77.3%) | 6,879 (81.6%) | 271 (46.3%) | 998 (65.3%) |  |
| Ex-smoker | 764 (7.2%) | 559 (6.6%) | 59 (10.1%) | 146 (9.5%) |  |
| Alcohol drinking |  |  |  |  | <0.001 |
| Current drinker | 2,354<br>(22.3%) | 1,664 (19.7%) | 235 (40.2%) | 455 (29.8%) |  |
| None drinker | 7,190<br>(68.2%) | 6,028 (71.5%) | 266 (45.5%) | 896 (58.6%) |  |
| Ex-drinker | 1,001 (9.5%) | 739 (8.8%) | 84 (14.4%) | 178 (11.6%) |  |
| Hours spent on exercise per day, mean (SD) | 1.2 (1.5) | 1.1 (1.4) | 2.0 (2.0) | 1.4 (1.8) | <0.001 |
| Physical activity ≥ 150 minutes/week | 6,232<br>(59.1%) | 4,901 (58.1%) | 432 (73.8%) | 899 (58.8%) | <0.001 |
| Self-reported overall health |  |  |  |  | <0.001 |
| Excellent | 4,211<br>(39.9%) | 3,388 (40.2%) | 305 (52.1%) | 518 (33.9%) |  |
| Very good | 4,235<br>(40.2%) | 3,533 (41.9%) | 146 (25.0%) | 556 (36.4%) |  |
| Good | 1,790<br>(17.0%) | 1,326 (15.7%) | 116 (19.8%) | 348 (22.8%) |  |
| Fair | 284 (2.7%) | 172 (2.0%) | 15 (2.6%) | 97 (6.3%) |  |
| Poor | 25 (0.2%) | 12 (0.1%) | 3 (0.5%) | 10 (0.7%) |  |
| Weight status |  |  |  |  | <0.001 |
| Underweight (BMI<18.5) | 1,093<br>(10.5%) | 878 (10.5%) | 77 (14.0%) | 138 (9.3%) |  |
| Normal (18.5<BMI<24) | 6,429<br>(61.8%) | 5,357 (64.0%) | 287 (52.2%) | 785 (53.0%) |  |
| Overweight (24<=BMI<28) | 2,108<br>(20.3%) | 1,574 (18.8%) | 131 (23.8%) | 403 (27.2%) |  |
| Obese (BMI>=28) | 768 (7.4%) | 559 (6.7%) | 55 (10.0%) | 154 (10.4%) |  |
| Number of chronic conditions |  |  |  |  | <0.001 |
| 0 | 8,431<br>(80.0%) | 8,431 (100.0%) | 0 (0.0%) | 0 (0.0%) |  |
| 1 | 1,024 (9.7%) | 0 (0.0%) | 119 (20.3%) | 905 (59.2%) |  |
| 2 | 591 (5.6%) | 0 (0.0%) | 203 (34.7%) | 388 (25.4%) |  |
| 3 | 307 (2.9%) | 0 (0.0%) | 142 (24.3%) | 165 (10.8%) |  |
| $\geq 4$ | 192 (1.8%) | 0 (0.0%) | 121 (20.7%) | 71 (4.6%) | |
| Hypertension | 732 (6.9%) | 0 (0.0%) | 146 (25.0%) | 586 (38.3%) | <0.001 |
| Heart problem | 437 (4.1%) | 0 (0.0%) | 134 (22.9%) | 303 (19.8%) | <0.001 |
| Depression | 270 (2.6%) | 0 (0.0%) | 29 (5.0%) | 241 (15.8%) | <0.001 |
| Food shortage | 2,837<br>(26.9%) | 1,914 (22.7%) | 350 (59.8%) | 573 (37.5%) | <0.001 |
| Medication shortage | 3,173<br>(30.1%) | 2,173 (25.8%) | 349 (59.7%) | 651 (42.6%) | <0.001 |

### Diabetes and self-reported COVID-19 infection in the family

Diabetes was associated with an increased risk of self-reported COVID-19 in the family. The prevalence of self-reported COVID-19 infection in the family were 5.2%, 37.9% and 18.6% among those without diabetes, with diabetes and with other NCDs, respectively. In a multivariable model, compared with those without diabetes, people with diabetes were 6.3 times (95% CI 5.07-7.84) more likely to report COVID-19 infection. There was no significant gender different in the association between diabetes and self-reported COVID-19 infection (**Figure 1**). However, the association between diabetes and COVID-19 infection was stronger in the city than in town and rural area (p-interaction 0.033). Among those perceiving themselves at high risk for COVD-19 infection, the association between diabetes and self-reported COVID-19 infection was lower than those who did not perceive high risk of COVID-19 (p-interaction <0.001). Among all the age groups, the association between diabetes and self-reported COVID-19 infection was the strongest in those aged 30-39 years.

**Figure 1.**
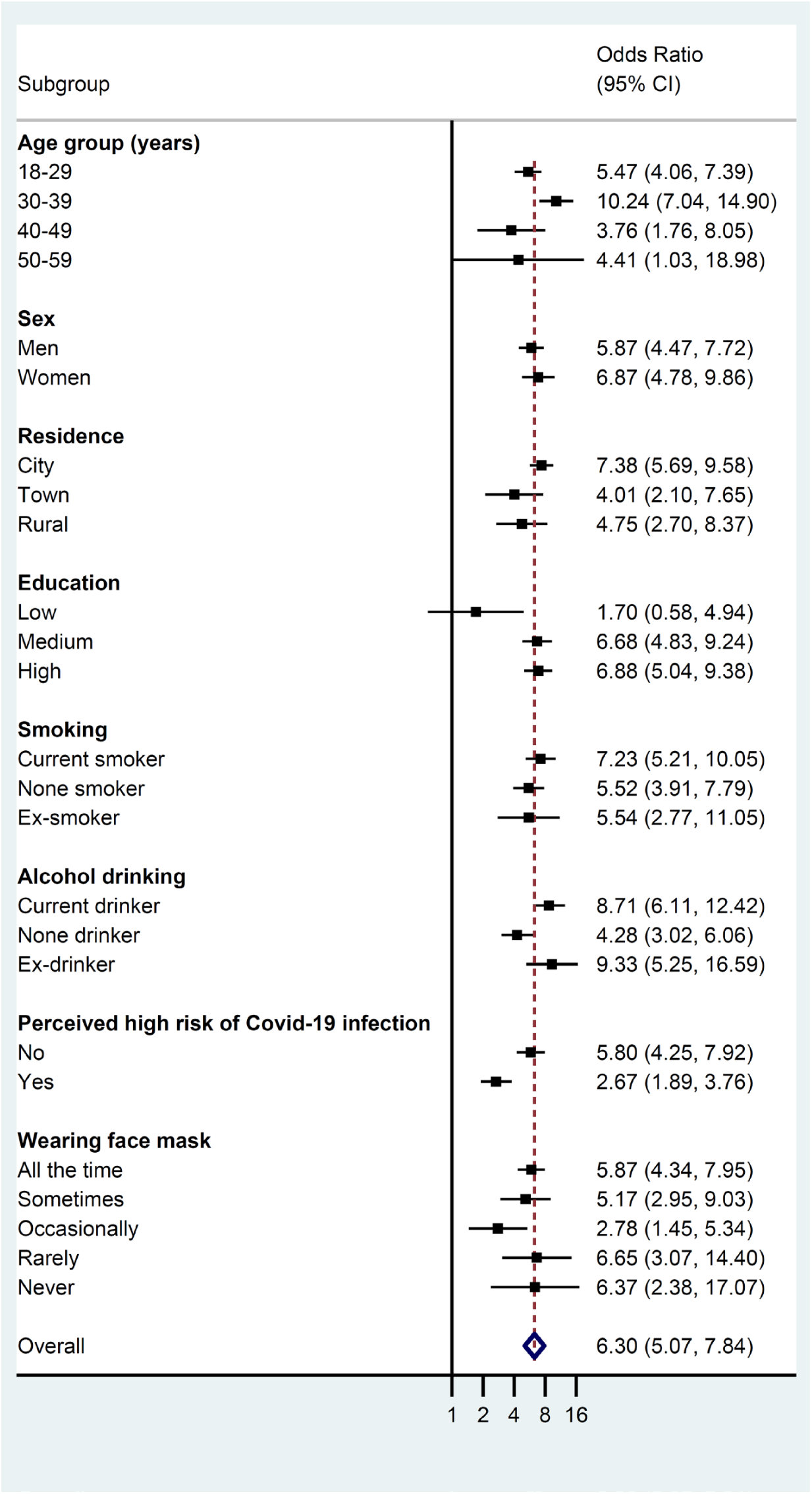
Associations between diabetes (as exposure variable) and self-reported COVID-19 infection (as outcome) by sociodemographic and lifestyle factors in The 2020 China COVID-19 Survey (N=10545) The models adjusted for age, gender, residence, education, smoking, alcohol drinking and physical activity (hours/day). Stratification variables were not adjusted in the corresponding models.

Among those with diabetes, medication shortage (PR 1.63 95%CI 1.20-2.22) and food shortage (PR 1.94 95%CI 1.33-2.84), and rarely wearing face mask (PR 2.12 95%CI 1.62-2.79) were positively associated with self-reported COVID-19 infection (**Figure 2**).

**Figure 2.**
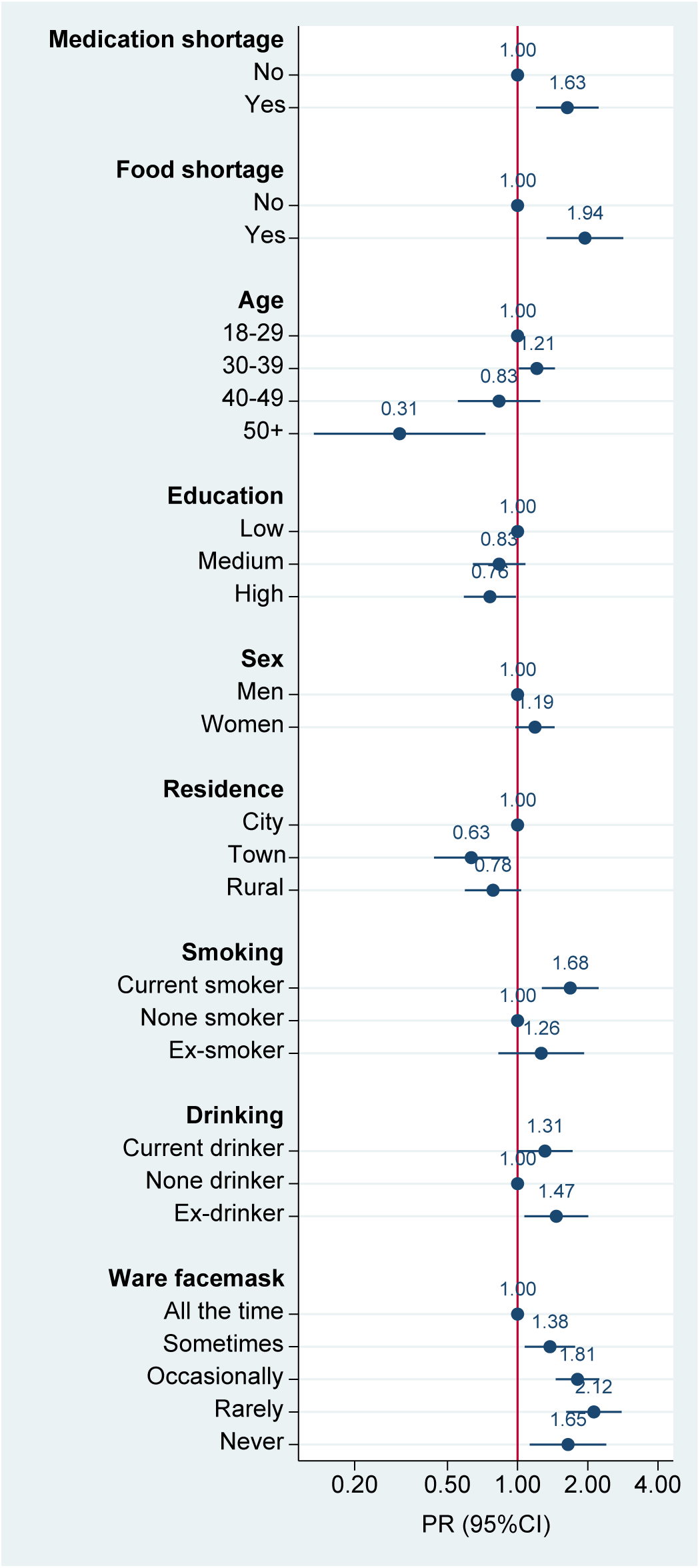
Factors associated with self-reported COVID-19 infection of participants or their family members among those with diabetes. All variables in the figure were mutually adjusted for in models. Multivariable Poisson regression with robust variance was used as the prevalence of COVID-19 infection was above 10%.

### Knowledge, attitudes, and practices towards COVID-19

Overall, people with diabetes were more likely to worry about being infected with COVID-19 than others, reporting themselves or family members as very likely to become infected (40.0% of respondents with diabetes vs 8.0% without) (**Table 2**). However, people with diabetes were less likely to correctly identify three symptoms of COVID-19 (59.3% among respondents with diabetes, 84.0% without diabetes and 69.9% with other NCDs).

**Table 2.** Perception and reaction to (i.e. behaviors and practice) the Covid19 pandemic among participants attending “The 2020 China COVID-19 Survey”, by their diabetic and non-communicable chronic disease (NCD) status (N=10545)

|  | **Total**<br>**N=10,545** | **Without**<br>**diabetes**<br>**N=8,431** | **With**<br>**diabetes**<br>**N=585** | **With other**<br>**NCD**<br>**N=1,529** | **p-**<br>**value** |
| --- | --- | --- | --- | --- | --- |
| 1.How serious of a public health threat do you think COVID-19 is or might become? |  |  |  |  | <0.001 |
| No threat at all | 686 (6.5%) | 402 (4.8%) | 111 (19.0%) | 173 (11.3%) |  |
| A little threat | 1,152 (10.9%) | 902 (10.7%) | 65 (11.1%) | 185 (12.1%) |  |
| Somewhat of a threat | 2,156 (20.4%) | 1,690 (20.0%) | 145 (24.8%) | 321 (21.0%) |  |
| A serious threat | 2,168 (20.6%) | 1,824 (21.6%) | 78 (13.3%) | 266 (17.4%) |  |
| A very serious threat | 4,383 (41.6%) | 3,613 (42.9%) | 186 (31.8%) | 584 (38.2%) |  |
| 2.How worried are you about being infected with COVID-19? |  |  |  |  | <0.001 |
| Very worried | 4,078 (38.7%) | 3,033 (36.0%) | 319 (54.5%) | 726 (47.5%) |  |
| Somewhat worried | 3,608 (34.2%) | 3,021 (35.8%) | 136 (23.2%) | 451 (29.5%) |  |
| A little worried | 2,375 (22.5%) | 1,997 (23.7%) | 110 (18.8%) | 268 (17.5%) |  |
| Not worried at all | 484 (4.6%) | 380 (4.5%) | 20 (3.4%) | 84 (5.5%) |  |
| 3.How likely do you think it is that you or someone in your family may get infected |  |  |  |  | <0.001 |
| Very likely | 1,237 (11.7%) | 673 (8.0%) | 234 (40.0%) | 330 (21.6%) |  |
| Somewhat likely | 2,573 (24.4%) | 1,972 (23.4%) | 158 (27.0%) | 443 (29.0%) |  |
| Not that likely | 4,711 (44.7%) | 4,065 (48.2%) | 135 (23.1%) | 511 (33.4%) |  |
| Not at all likely | 2,024 (19.2%) | 1,721 (20.4%) | 58 (9.9%) | 245 (16.0%) |  |
| 4.Correctly identified 3 symptoms of the coronavirus | 8,501 (80.6%) | 7,085 (84.0%) | 347 (59.3%) | 1,069 (69.9%) | <0.001 |
| 5.Correctly identified 3 prevention methods of the coronavirus | 9,219 (87.4%) | 7,675 (91.0%) | 375 (64.1%) | 1,169 (76.5%) | <0.001 |
| 5.Always practiced four prevention measures of the coronavirus | 6,507 (61.7%) | 5,496 (65.2%) | 261 (44.6%) | 750 (49.1%) | <0.001 |
| 6.Strongly agreed with 5 community prevention methods of the coronavirus | 6,445 (61.1%) | 5,441 (64.5%) | 245 (41.9%) | 759 (49.6%) | <0.001 |

People with diabetes were also less likely to consistently practice COVID-19-preventive measures (**Supplement Figure 1**). Fewer than 60% of them consistently practiced the four cardinal preventive measures (wearing face masks, frequent hand washing, avoiding unnecessary outdoor activity, and avoiding social gatherings). In contrast, more than 70% of those without diabetes reported practicing these four preventive measures “all the time.”

### Management of diabetes and other NCDs during COVID-19 pandemic

Half of those with diabetes reported current blood glucose levels to be above the normal range (**Supplement Table 1**), 31.6% had their last glucose test more than 6 months ago (**Table 3**). A total of 28.2% of those with diabetes reported daily measurement of blood pressure, but 71.3% of them reported that their routine medical consultation was affected due to COVID-19, and 21.5% were tested for blood lipids one year ago.

**Table 3.** Management of diabetes and other and non-communicable chronic disease (NCD) status among participants attending “The 2020 China COVID 19 Survey”, by diabetic and non-communicable chronic disease (NCD) status (N=10,545)

|  | **Total**<br>**N=10,545** | **Without**<br>**diabetes**<br>**N=8,431** | **With**<br>**diabetes**<br>**N=585** | **With other**<br>**NCD**<br>**N=1,529** | **p-**<br>**value** |
| --- | --- | --- | --- | --- | --- |
| 1. Has your routine medical care or medical consultation been affected due to COVID |  |  |  |  | <0.001 |
| Yes | 4,911 (46.6%) | 3,624 (43.0%) | 417 (71.3%) | 870 (56.9%) |  |
| No | 5,634 (53.4%) | 4,807 (57.0%) | 168 (28.7%) | 659 (43.1%) |  |
| 2. Have you visited the hospital due to suspected COVID-19 infection? |  |  |  |  | <0.001 |
| Yes | 1,742 (16.5%) | 992 (11.8%) | 304 (52.0%) | 446 (29.2%) |  |
| No | 8,803 (83.5%) | 7,439 (88.2%) | 281 (48.0%) | 1,083 (70.8%) |  |
| 3. When was the last time you got blood pressure measured? |  |  |  |  | <0.001 |
| Daily measurement | 1,245 (11.8%) | 789 (9.4%) | 165 (28.2%) | 291 (19.0%) |  |
| Within 7 days | 1,596 (15.1%) | 1,167 (13.8%) | 96 (16.4%) | 333 (21.8%) |  |
| Within 1 month | 2,016 (19.1%) | 1,487 (17.6%) | 188 (32.1%) | 341 (22.3%) |  |
| Within 3 months | 1,424 (13.5%) | 1,135 (13.5%) | 74 (12.6%) | 215 (14.1%) |  |
| 3 months ago | 2,413 (22.9%) | 2,167 (25.7%) | 40 (6.8%) | 206 (13.5%) |  |
| Never | 1,851 (17.6%) | 1,686 (20.0%) | 22 (3.8%) | 143 (9.4%) |  |
| 4. When was the last time you got blood sugar measured? |  |  |  |  | <0.001 |
| Within 6 months | 3,678 (34.9%) | 2,720 (32.3%) | 358 (61.2%) | 600 (39.2%) |  |
| Within 1 year | 2,656 (25.2%) | 2,154 (25.5%) | 97 (16.6%) | 405 (26.5%) |  |
| 1 year ago | 1,509 (14.3%) | 1,176 (13.9%) | 88 (15.0%) | 245 (16.0%) |  |
| Never | 2,702 (25.6%) | 2,381 (28.2%) | 42 (7.2%) | 279 (18.2%) |  |
| 5. When was the last time you got blood lipids measured? |  |  |  |  | <0.001 |
| Within 6 months | 3,017 (28.6%) | 2,203 (26.1%) | 314 (53.7%) | 500 (32.7%) |  |
| Within 1 year | 2,999 (28.4%) | 2,392 (28.4%) | 126 (21.5%) | 481 (31.5%) |  |
| 1 year ago | 1,398 (13.3%) | 1,037 (12.3%) | 92 (15.7%) | 269 (17.6%) |  |
| Never measured blood lipids | 3,131 (29.7%) | 2,799 (33.2%) | 53 (9.1%) | 279 (18.2%) |  |

### COVID-19 related health outcomes

People with diabetes were more likely to report their health was significantly worse (33.7% vs. 7.2%) and have a high worry index because of the pandemic than those without diabetes (**Supplement Table 1**). In a multivariable model, the PR for significant health worsening during the pandemic was 2.69 (95% CI 2.31-3.14) for those with diabetes compared with those without diabetes (**Supplement Table 2**).

## DISCUSSION

This paper is the first from examining the association between diabetes and COVID-19 and how the risk of infection can be mitigated by preventive measures such as face mask wearing. The findings highlight important lessons for dealing with future pandemics of this magnitude. Here, diabetes was associated with a six-fold increased risk of reported COVID-19 infection for respondents or their family members. Given the highly infectious nature of the virus and its well-documented high community transmission rate, this was an expected outcome of a family cluster.^18^ Compared to those who always wore a face mask in public settings, people with diabetes who opted to rarely wear a face mask had a two-fold increased risk of suspected COVID-19 infection.

The benefit of wearing a face mask in the community to reduce the risk of transmission of COVID-19 has yet to be established, with conflicting evidence and views.^19,20^ To date, the efficacy of community-wide masking during the COVID-19 pandemic has not been clearly investigated. Studies found that up to 80% of people with COVID-19 were mild or asymptomatic.^21^ Thus subclinical or asymptomatic SARS-CoV-2 patients may play an important role in the perpetuation of the pandemic. To reduce the presence of viral particles in droplets and aerosol generated by symptomatic or asymptomatic SARS-CoV-2–infected individuals, Leung et al. suggested the need for universal use of cloth face covering or surgical masks when available. ^22^

Our findings of a two-fold increase in risk of COVID-19 infection for those not using face masks in public support their use. This is especially important if social distancing is not possible or is difficult to achieve. Donning masks is recommended by the WHO, the US Centers for Disease Control and Prevention,^23^ and Chu and colleagues’ recent review ^24^ wherein they reported that face masks reduced COVID-19 infection risk in healthcare settings. Increasingly, public health messaging maintains that masks should be worn universally in public settings, such as large crowds, indoor settings (grocery stores or supermarkets), and while using public transportation.

Another important finding of our study was that people with diabetes had limited knowledge about preventive measures for COVID-19, even though they perceived they were at a higher risk of infection (40.0% in diabetes and 8.0% without diabetes). Up to 55% of people with diabetes always practiced preventive measures, including wearing face masks. A growing body of clinical and population studies shows that patients with diabetes are more likely to develop severe symptoms and complications of COVID-19 infection,^25,26^ and they have much higher admission rates in intensive care units. As more countries recognize the serious health outcomes of COVID-19 infection, including increased risk of death in people with diabetes and other non-communicable diseases, these individuals should clearly be listed as at a higher risk in national media and other public health messages.

Despite the widely-held view that the Chinese central and local governments made efforts to meet citizens’ daily living needs during the national lockdown, we found that nearly 60% of adults with diabetes reported experiencing food (59.8%) or medication shortages (59.7%) during the pandemic. This was much higher than those without diabetes (22.7 % and 25.8%, respectively). The social, economic, and political implications are concerning. A Danish study^10^ conducted during the pandemic also reported that about 25% of people with diabetes feared medical supply shortages and 10% reported food shortage. This was in a country with universal health coverage and a highly reputable social welfare system in place. Our surveyed population with diabetes had higher income but poorer levels of educational attainment compared to those without diabetes. Thus, income was likely not the barrier to securing food and medicine. Some plausible explanations are that a) people with diabetes remained at home (for fear of being infected and a lack of knowledge about protective measures) and therefore could not easily access or procure medicine and food; and b) since people with diabetes depend more on specific food items and medication to control their diabetes, they are more concerned about these areas. Given the self-reported nature of the data, it is also possible that in some cases there was more of a perceived shortage of food and medication than an actual shortage.

Among people with diabetes, those who reported experiencing medication shortages had a 63% higher rate of self-reported COVID-19 infection. It is now well-established that poor metabolic control places people with diabetes at the highest risk for COVID-19 infection^27^ along with its associated devastating morbidity and mortality.^25^ Almost half of survey respondents with diabetes had not monitored their blood glucose levels for over six months. This links directly with the previous statement about optimal metabolic control of diabetes being protective against serious consequences of COVID-19 infection.^27^ Certainly, the pandemic has caused a disruption of the regular medical care and contact normally received by many people with diabetes, not to mention the regular monitoring of their glucose control. This poor compliance may be partly stress-induced, though it is just as likely due to the inability to access care during the pandemic. Nevertheless, as we know that poor metabolic control heightens the risk and severity of a COVID-19 infection, this is a matter of public health concern that needs to be acted on now and considered in future pandemics.

Finally, half of the respondents with diabetes visited the hospital for testing due to suspected COVID-19 infection. Presumably, these people were aware of the dangers that COVID-19 posed to their health. Further, this may reflect COVID-19-related anxiety in people with diabetes, since their prevalence of COVID-19 in China ranged from 4-22%.^2^

This study has several key strengths. First, we collected comprehensive data related to COVID-19 and NCDs including T2DM. This allowed examination of the interesting and complex relationships among them. Second, to our knowledge, no other studies have reported findings on the associations between diabetes and COVID-19 on a large sample in China, where COVID-19 was first reported, and government-imposed measures were introduced to control its spread. Our findings can shed light on other populations at heightened risk of COVID-19 infection in an environment rife with exposure to risk. In addition, given the survey’s anonymous design, such an approach can collect people’s honest views and sensitive information. It is also a more feasible data collection method in special situations like COVID-19, where face-to-face interviews are difficult and risky.

However, the study also has limitations. First, the potential for self-selection bias is recognized in such surveys. However, it is somewhat reassuring that the prevalence of self-reported diabetes and other NCDs in our data is similar to other population surveys in China.^28,29^ Second, the sample was relatively younger than the national average. Therefore, findings may not be generalizable to the entire population. Third, the survey questionnaire did not ask participants to identify the specific type of diabetes that they had been diagnosed, though it is well documented that the vast majority of diabetes cases in China are T2DM.^30^

In conclusion, people with diabetes and their family members were found to have an increased risk of COVID-19 infection, and were more like to report shortages of food and medications during the national COVID-19 lockdown in China. Those with diabetes exhibited a variety of characteristics and behaviors which would be expected to increase both the risk of COVID-19 infection and poorer outcomes once infected. Our findings suggest that promoting public health education, better monitoring of health parameters, and stricter adherence to recommended preventive measures are particularly important among adults with diabetes. With the expanding COVID-19 pandemic, there is also an urgent need to implement measures to address anxiety management and provide practical guidance and emotional support to those at greater risk, in particular people with diabetes and possibly other NCDs.

## Data Availability

Data is only available upon request and approval.

## Acknowledgements

We thank all the study participants and collaborators and staff members who have contributed to the study. We thank Guorui Ruan, Lihua Yan, and Bingtong Zhao for their special assistance supporting this project.

## Author contributions

YW, ZS and AF contributed to the study design and drafting the manuscript. ZS, AF and YW contributed to the data analysis and drafted the manuscript. YW directed data collection and provided administrative support for the project. All authors contributed to interpretation of the data, revised the report, and approved the final version for publication.

---

**Supplement Table 1.** Effects of Covid-19 on health outcomes among participants attending The 2020 China COVID-19 Survey, by diabetes status (N=10545)

|  | **Total**<br>**N=10,545** | **None**<br>**N=8,431** | **Prevalence**<br>**Diabetes**<br>**N=585** | **Other NCD**<br>**N=1,529** | **p-value** |
| --- | --- | --- | --- | --- | --- |
| Self-reported overall health |  |  |  |  | <0.001 |
| Excellent | 4,211 (39.9%) | 3,388 (40.2%) | 305 (52.1%) | 518 (33.9%) |  |
| Very good | 4,235 (40.2%) | 3,533 (41.9%) | 146 (25.0%) | 556 (36.4%) |  |
| Good | 1,790 (17.0%) | 1,326 (15.7%) | 116 (19.8%) | 348 (22.8%) |  |
| Fair | 284 (2.7%) | 172 (2.0%) | 15 (2.6%) | 97 (6.3%) |  |
| Poor | 25 (0.2%) | 12 (0.1%) | 3 (0.5%) | 10 (0.7%) |  |
| How has COVID-19 affected your health? |  |  |  |  | <0.001 |
| Significantly worse | 1,096 (10.4%) | 609 (7.2%) | 197 (33.7%) | 290 (19.0%) |  |
| Slightly worse | 1,828 (17.3%) | 1,327 (15.7%) | 126 (21.5%) | 375 (24.5%) |  |
| Slightly better | 1,317 (12.5%) | 971 (11.5%) | 112 (19.1%) | 234 (15.3%) |  |
| Significantly better | 601 (5.7%) | 445 (5.3%) | 34 (5.8%) | 122 (8.0%) |  |
| No change | 5,703 (54.1%) | 5,079 (60.2%) | 116 (19.8%) | 508 (33.2%) |  |
| Has your routine medical care or medical consultation been affected due to COVID |  |  |  |  | <0.001 |
| Yes | 4,911 (46.6%) | 3,624 (43.0%) | 417 (71.3%) | 870 (56.9%) |  |
| No | 5,634 (53.4%) | 4,807 (57.0%) | 168 (28.7%) | 659 (43.1%) |  |
| Have you visited the hospital due to suspected COVID-19 infection? |  |  |  |  | <0.001 |
| Yes | 1,742 (16.5%) | 992 (11.8%) | 304 (52.0%) | 446 (29.2%) |  |
| No | 8,803 (83.5%) | 7,439 (88.2%) | 281 (48.0%) | 1,083 (70.8%) |  |
| What do you think of your current blood pressure level? |  |  |  |  | <0.001 |
| Above normal range | 1,478 (14.0%) | 758 (9.0%) | 274 (46.8%) | 446 (29.2%) |  |
| Within normal range | 7,251 (68.8%) | 6,213 (73.7%) | 213 (36.4%) | 825 (54.0%) |  |
| Below normal range | 427 (4.0%) | 222 (2.6%) | 81 (13.8%) | 124 (8.1%) |  |
| Don't know | 1,389 (13.2%) | 1,238 (14.7%) | 17 (2.9%) | 134 (8.8%) |  |
| What do you think of your current blood sugar level? |  |  |  |  | <0.001 |
| Above normal range | 1,214 (11.5%) | 618 (7.3%) | 293 (50.1%) | 303 (19.8%) |  |
| Within normal range | 6,646 (63.0%) | 5,582 (66.2%) | 194 (33.2%) | 870 (56.9%) |  |
| Below normal range | 307 (2.9%) | 126 (1.5%) | 67 (11.5%) | 114 (7.5%) |  |
| Don't know | 2,378 (22.6%) | 2,105 (25.0%) | 31 (5.3%) | 242 (15.8%) |  |
| What do you think of your current blood lipid profile? |  |  |  |  | <0.001 |
| Normal | 7,310 (69.3%) | 5,858 (69.5%) | 440 (75.2%) | 1,012 (66.2%) |  |
| Abnormal | 615 (5.8%) | 292 (3.5%) | 87 (14.9%) | 236 (15.4%) |  |
| Don't know | 2,620 (24.8%) | 2,281 (27.1%) | 58 (9.9%) | 281 (18.4%) |  |
| How is your current weight compared to your weight before COVID-19? |  |  |  |  | <0.001 |
| Unchanged (increased or decreased within 1kg) | 4,271 (40.5%) | 3,583 (42.5%) | 207 (35.4%) | 481 (31.5%) |  |
| Increased by 1-2.5kg | 3,486 (33.1%) | 2,708 (32.1%) | 204 (34.9%) | 574 (37.5%) |  |
| Increased by 2.5kg or more | 1,800 (17.1%) | 1,357 (16.1%) | 123 (21.0%) | 320 (20.9%) |  |
| Decreased by 1-2.5kg | 594 (5.6%) | 454 (5.4%) | 38 (6.5%) | 102 (6.7%) |  |
| Decreased by >2.5kg | 241 (2.3%) | 190 (2.3%) | 11 (1.9%) | 40 (2.6%) |  |
| Don't know | 153 (1.5%) | 139 (1.6%) | 2 (0.3%) | 12 (0.8%) |  |
| Worries index | 0.0 (1.0) | -0.1 (0.9) | 0.6 (1.2) | 0.4 (1.1) | <0.001 |
| Self-reported COVID-19 infection or family member infection | 946 (9.0%) | 439 (5.2%) | 222 (37.9%) | 285 (18.6%) | <0.001 |
| Self-reported COVID-19 infection or family member infection (strict definition)<sup>a</sup> | 647 (6.1%) | 270 (3.2%) | 186 (31.8%) | 191 (12.5%) | <0.001 |

Data were presented as mean (SD) for continuous measures, and n (%) for categorical measures.
<sup>a</sup> A positive answer to all three questions related to COVID-19 infection including visited hospital and felt discrimination.

**Supplement Table 2.** Association between diabetes and Covid19 infection, self-reported health worsening, worry index among participants attending The China 2020 COVID-19 Survey (N=10545)

|  | Without diabetes | Diabetes | With other chronic diseases |
| --- | --- | --- | --- |
| Self-reported suspected Covid19 infection |  |  |  |
| Prevalence (%) | 5.2 | 38.0 | 18.6 |
| OR (95%CI) | 1.00 | 6.30 (5.07-7.84) | 3.37 (2.83-4.01) |
| Significant health worsening during pandemic |  |  |  |
| Prevalence (%) | 7.2 | 33.7 | 19.0 |
| PR (95%CI) | 1.00 | 2.69 (2.31-3.14) | 2.09 (1.82-2.39) |
| Worry index |  |  |  |
| Mean (SD) | -0.12(0.93) | 0.56(1.21) | 0.44(1.09) |
| B (95%CI) | 0.00 | 0.58 (0.50-0.66) | 0.55 (0.50-0.61) |

To estimate PRs and 95%CIs, all models adjusted for age, education, gender, residence, smoking, drinking, and face mask wearing.

**Supplement Figure 1.**
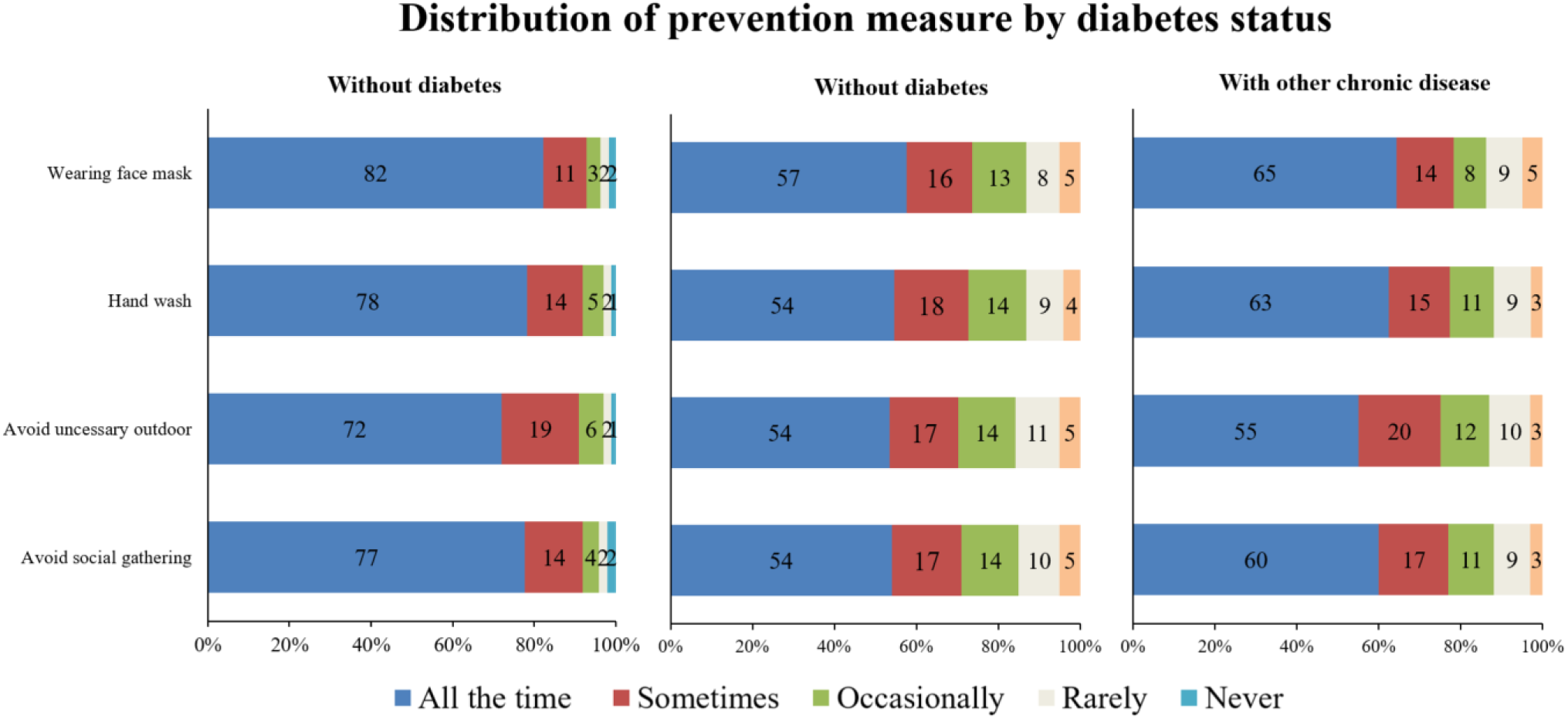
Distribution of COVID-19 preventive measures among participants attending The 2020 China COVID-19 Survey, by diabetes status.

## REFERENCES

1. Yang J, Zheng Y, Gou X, et al. Prevalence of comorbidities in the novel Wuhan coronavirus (COVID-19) infection: a systematic review and meta-analysis. Int J Infect Dis 2020.

2. Hussain A, Bhowmik B, do Vale Moreira NC. COVID-19 and diabetes: Knowledge in progress. Diabetes Res Clin Pract 2020; 162: 108142.

3. Gamble A, Pham Q, Goyal S, Cafazzo JA. The Challenges of COVID-19 for People Living With Diabetes: Considerations for Digital Health. JMIR Diabetes 2020; 5(2): e19581.

4. Schofield J, Leelarathna L, Thabit H. COVID-19: Impact of and on Diabetes. Diabetes Ther 2020.

5. Rosenbaum L. The Untold Toll — The Pandemic’s Effects on Patients without Covid-19. New England Journal of Medicine 2020.

6. Beran D, Aebischer Perone S, Castellsague Perolini M, et al. Beyond the virus: Ensuring continuity of care for people with diabetes during COVID-19. Prim Care Diabetes 2020.

7. Saeedi P, Petersohn I, Salpea P, et al. Global and regional diabetes prevalence estimates for 2019 and projections for 2030 and 2045: Results from the International Diabetes Federation Diabetes Atlas, 9(th) edition. Diabetes Res Clin Pract 2019; 157: 107843.

8. Ma QX, Shan H, Zhang HL, Li GM, Yang RM, Chen JM. Potential utilities of mask-wearing and instant hand hygiene for fighting SARS-CoV-2. Journal of medical virology 2020.

9. Zhong BL, Luo W, Li HM, et al. Knowledge, attitudes, and practices towards COVID-19 among Chinese residents during the rapid rise period of the COVID-19 outbreak: a quick online cross-sectional survey. Int J Biol Sci 2020; 16(10): 1745–52.

10. Joensen LE, Madsen KP, Holm L, et al. Diabetes and COVID-19: psychosocial consequences of the COVID-19 pandemic in people with diabetes in Denmark-what characterizes people with high levels of COVID-19-related worries? Diabet Med 2020; 37(7): 1146–54.

11. Iqbal M. WeChat Revenue and Usage Statistics 2020. https://www.businessofapps.com/data/wechat-statistics/ (accessed June 27 2020).

12. Wilkins KC, Lang AJ, Norman SB. Synthesis of the psychometric properties of the PTSD checklist (PCL) military, civilian, and specific versions. Depress Anxiety 2011; 28(7): 596–606.

13. Wolf MS, Serper M, Opsasnick L, et al. Awareness, Attitudes, and Actions Related to COVID-19 Among Adults With Chronic Conditions at the Onset of the U.S. Outbreak. Annals of Internal Medicine 2020; 0(0): null.

14. Conway LG, Woodard Shailee R., Zubrod A. Social Psychological Measurements of COVID-19: Coronavirus Perceived Threat, Government Response, Impacts, and Experiences Questionnaires. 2020. https://www.nlm.nih.gov/dr2/Scales_from_Social_Psychological_Measurements_of_COVID-19.pdf (accessed April 26 2020).

15. Lee PH, Macfarlane DJ, Lam TH, Stewart SM. Validity of the International Physical Activity Questionnaire Short Form (IPAQ-SF): a systematic review. Int J Behav Nutr Phys Act 2011; 8: 115.

16. Hays RD, Spritzer KL, Thompson WW, Cella D. U.S. General Population Estimate for “Excellent” to “Poor” Self-Rated Health Item. J Gen Intern Med 2015; 30(10): 1511–6.

17. Barros AJD, Hirakata VN. Alternatives for logistic regression in cross-sectional studies: an empirical comparison of models that directly estimate the prevalence ratio. BMC Medical Research Methodology 2003; 3(1): 21.

18. Bi Q, Wu Y, Mei S, et al. Epidemiology and transmission of COVID-19 in 391 cases and 1286 of their close contacts in Shenzhen, China: a retrospective cohort study. The Lancet Infectious Diseases.

19. Cheng VC-C, Wong S-C, Chuang VW-M, et al. The role of community-wide wearing of face mask for control of coronavirus disease 2019 (COVID-19) epidemic due to SARS-CoV-2. J Infect 2020; 81(1): 107–14.

20. Esposito S, Principi N. To mask or not to mask children to overcome COVID-19. Eur J Pediatr 2020: 1–4.

21. Day M. Covid-19: four fifths of cases are asymptomatic, China figures indicate. BMJ 2020; 369: m1375.

22. Leung NHL, Chu DKW, Shiu EYC, et al. Respiratory virus shedding in exhaled breath and efficacy of face masks. Nature Medicine 2020; 26(5): 676–80.

23. Centers for Disease Control and Prevention. Coronavirus disease 2019 (COVID-19). How to protect yourself & others. 2020. https://www.cdc.gov/coronavirus/2019-ncov/prevent-getting-sick/prevention.html (accessed June 25 2020).

24. Chu DK, Akl EA, Duda S, et al. Physical distancing, face masks, and eye protection to prevent person-to-person transmission of SARS-CoV-2 and COVID-19: a systematic review and meta-analysis. The Lancet.

25. Fang L, Karakiulakis G, Roth M. Are patients with hypertension and diabetes mellitus at increased risk for COVID-19 infection? Lancet Resp Med 2020; 8(4): E21–E.

26. Zhang Y, Li H, Zhang J, et al. The Clinical Characteristics and Outcomes of Diabetes Mellitus and Secondary Hyperglycaemia Patients with Coronavirus Disease 2019: a Single-center, Retrospective, Observational Study in Wuhan. Diabetes Obes Metab 2020.

27. Bornstein SR, Dalan R, Hopkins D, Mingrone G, Boehm BO. Endocrine and metabolic link to coronavirus infection. Nat Rev Endocrinol 2020; 16(6): 297–8.

28. Wang L, Gao P, Zhang M, et al. Prevalence and Ethnic Pattern of Diabetes and Prediabetes in China in 2013. JAMA 2017; 317(24): 2515–23.

29. Wang Y, Wang L, Qu W. New national data show alarming increase in obesity and noncommunicable chronic diseases in China. Eur J Clin Nutr 2017; 71(1): 149–50.

30. Hu C, Jia W. Diabetes in China: Epidemiology and Genetic Risk Factors and Their Clinical Utility in Personalized Medication. Diabetes 2018; 67(1): 3–11.

